# Establishing a Core Outcome Set for Creatine Transporter Deficiency and Guanidinoacetate Methyltransferase Deficiency

**DOI:** 10.1101/2024.09.06.24313213

**Authors:** Zahra Nasseri Moghaddam, Emily K. Reinhardt, Audrey Thurm, Beth K. Potter, Maureen Smith, Celeste Graham, Beth H. Tiller, Steven A. Baker, Deborah A. Bilder, Regina Bogar, Jacobus Britz, Rachel Cafferty, Daniel P. Coller, Ton J. DeGrauw, Vicky Hall, Gerald S. Lipshutz, Nicola Longo, Saadet Mercimek-Andrews, Judith S. Miller, Marzia Pasquali, Gajja S. Salomons, Andreas Schulze, Celine P. Wheaton, Kayla F. Williams, Sarah P. Young, Jasmine Li, Sofia Balog, Theresa Selucky, Sylvia Stockler-Ipsiroglu, Heidi Wallis

## Abstract

Creatine transporter (CTD) and guanidinoacetate methyltransferase (GAMT) deficiencies are rare inborn errors of creatine metabolism, resulting in cerebral creatine deficiency. Patients commonly exhibit intellectual and developmental disabilities, often accompanied by behavior problems, delayed speech, seizures, and motor impairments. There is currently no efficacious treatment for CTD, while the current management for GAMT requires lifelong treatment with a protein restricted diet and intake of high amounts of oral supplements. Efforts to develop effective, sustainable treatments for these disorders are limited by the lack of clinical and patient-derived meaningful outcomes. A core outcome set (COS) can facilitate consensus about outcomes for inclusion in studies. Unfortunately, patient and caregiver perspectives have historically been overlooked in the COS development process, thus limiting their input into the outcome selection. We partnered with caregivers and health professionals to establish the first COS for CTD and GAMT. The COS developed includes seven outcomes (“Adaptive Functioning”, “Cognitive Functioning”, “Emotional Dysregulation”, “MRS Brain Creatine”, “Seizure/Convulsions”, “Expressive Communication”, and “Fine Motor Functions”) for both CTD and GAMT, and an additional outcome for GAMT (“Serum/Plasma Guanidinoacetate”) that are important to stakeholders and consequently should be considered for measurement in every clinical trial. Caregivers were valued partners throughout the COS development process, which increased community engagement and facilitated caregiver empowerment. We expect this COS will ensure a patient-centered approach for accelerating drug development for CTD and GAMT, make clinical trial results comparable, minimize bias in clinical trial outcome selection, and promote efficient use of resources.

**1-sentence take home message:** A core outcome set for creatine transporter (CTD) and guanidinoacetate methyltransferase (GAMT) deficiencies was created through a multiphase process in partnership with caregivers and health professionals.

## INTRODUCTION

Creatine transporter (CTD, MIM:300352) and guanidinoacetate methyltransferase (GAMT, MIM:612736) deficiencies are rare inborn errors of creatine metabolism and transport, resulting in cerebral creatine deficiency.^1–3^ CTD is an X-linked disorder caused by hemizygous or heterozygous pathogenic variants in *SLC6A8*, disrupting the cellular uptake of creatine in the brain.^3–4^ GAMT is an autosomal recessive disorder caused by biallelic pathogenic variants in *GAMT*, preventing the biosynthesis of creatine.^5^ GAMT also causes an accumulation of guanidinoacetate (GAA), which is neurotoxic.^5–9^ Similar to other genetic conditions associated with neurodevelopmental disorders, individuals with CTD and GAMT commonly exhibit intellectual and/or developmental disabilities, often accompanied by behavior problems, delayed speech, seizures, and motor impairments.^5,10–12^

There is currently no efficacious disease-modifying treatment for CTD. However, some evidence suggests that creatine oral supplementation and creatine precursors, arginine and glycine, partially improve symptoms in some CTD patients.^4,13–15^ CTD patients currently utilize medications and therapies (e.g., speech, occupational) to improve functioning. For GAMT, oral creatine supplementation has shown efficacy in partially restoring brain creatine levels while dietary arginine restriction and L-ornithine supplementation may further reduce GAA levels.^16–21^ The current management for GAMT requires lifelong treatment with a protein restricted diet and intake of high amounts of supplements.^18, 22–23^ In both CTD and GAMT, timely diagnosis and early intervention have been associated with enhanced quality of life and reduced cognitive impairment.^2,4,17,24^ Worldwide efforts are ongoing to understand the long-term impacts of CTD and GAMT.^3,25–28^

Implementing clinical trials in rare diseases such as CTD and GAMT is especially challenging due to factors that include a small population and substantial heterogeneity in phenotypes.^29–33^ Identifying outcomes (e.g., clinical, behavioral, and/or laboratory-based) that assess treatment efficacy is critical to clinical trial success. Unfortunately, measuring different outcomes between trials makes it difficult to compare the efficacy of different therapeutics, especially when treatments target disease-modification of multi-systemic conditions affecting development. ^34–35^ Historically, trial outcomes have been selected without input from patients and caregivers.^36–37^ Patients and caregivers are often the best equipped to identify the meaningful changes that would improve daily functioning and overall health as a result of a treatment. The U.S. Food and Drug Administration (FDA) and other organizations now require patient and caregiver input as part of the clinical trial design process.^38^

A core outcome set (COS) can be used to address these obstacles to rare disease research and inform trial design. A COS is a small set of outcomes that are established as important to be collected in every research study of the same disease.^39–40^ A COS promotes efficient use of resources and minimizes risk of bias by incorporating stakeholder perspectives, thereby ensuring that the evidence generated addresses outcomes that reflect meaningful treatment responses.^41^ It also facilitates uniformity in the selection, measurement, and reporting of outcomes to simplify comparison across studies.^41–42^ To support future clinical trials, we established the first COS for CTD and GAMT.

## METHODS

### Project Design

We relied on the Core Outcome Measures in Effectiveness Trials (COMET) initiative to guide us in developing our COS.^41,43–44^ Our findings are reported in accordance with the COS Standard for Reporting statement (Supplemental 1).^45^

To ensure that the final COS reflected the patient and caregiver perspective, active patient and caregiver engagement was a key design element throughout the COS development process. The project was conducted in three sequential phases: 1) Candidate Outcome Selection, 2) Delphi Surveys, 3) Consensus Workshop (Figure 1).

**Figure 1.**
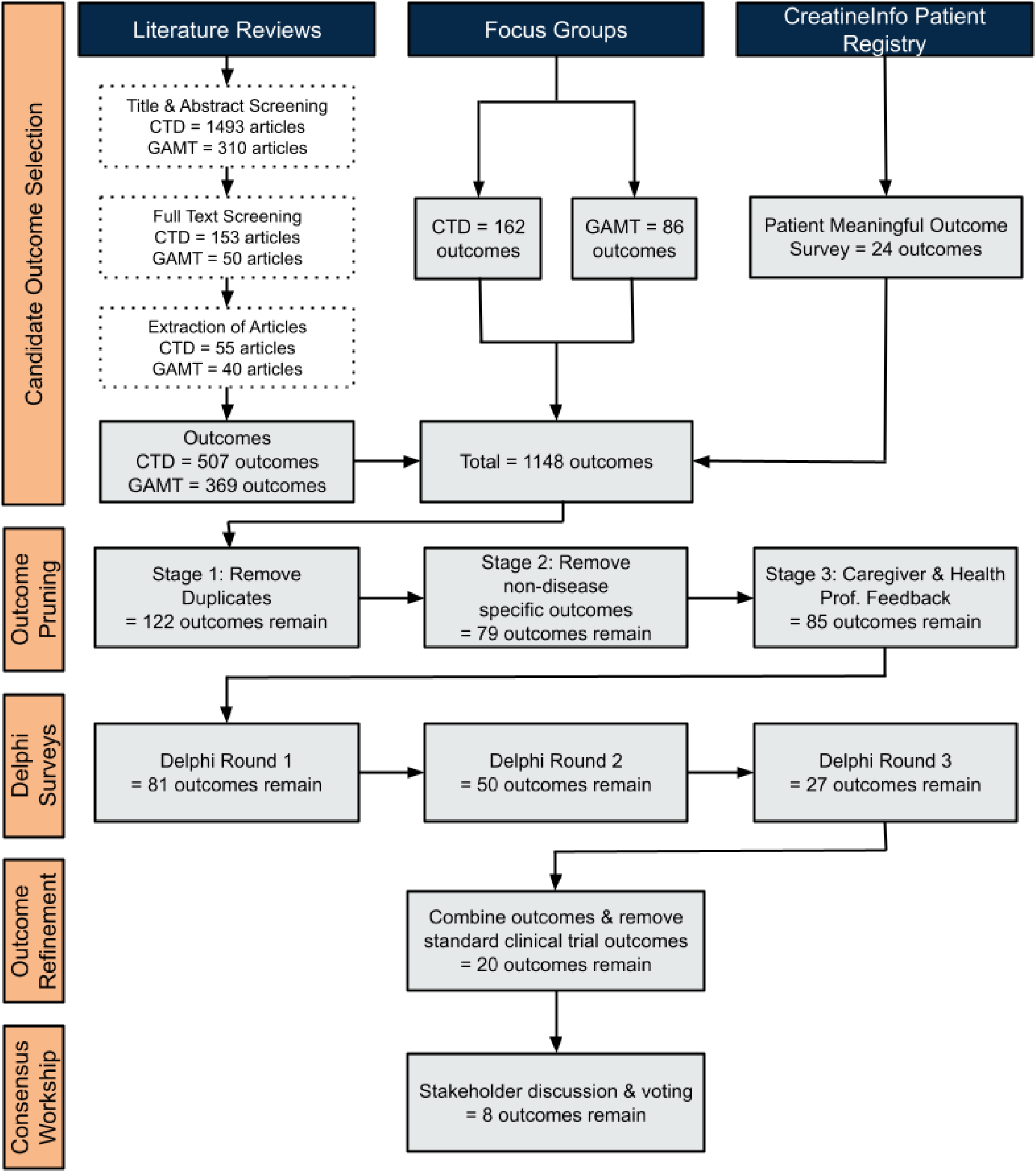
Outcome selection process for COS development for CTD and GAMT. During the candidate outcome selection phase, the total number of candidate outcomes gathered during each stage are identified. During the remaining project phases, the total number of remaining outcomes at the end of each stage are identified.

### Patient and Caregiver Engagement

CTD and GAMT result in severe neurodevelopmental disorders, with affected individuals often nonverbal. Since caregivers must therefore serve as the patient’s voice, caregivers were critical in representing lived experiences. An active partnership between caregivers, clinicians, researchers, and scientists was utilized. This project was a component of a broader Patient-Centered Outcomes Research Institute (PCORI)-funded patient engagement project “Parents Advancing REsearch NeTworkS” or PAReNts.^46^ PAReNts aimed to empower caregivers and build their capacity to partner with researchers. Broadly, members of the PAReNts project received training and education about clinical trial research and COS development, shared their personal experiences, participated in focus groups, and actively engaged with other stakeholders to help develop the final COS for CTD and GAMT.

### Candidate Outcome Selection

The goal of the candidate outcome selection phase was to identify possible outcomes for CTD and GAMT from a variety of sources and diverse perspectives.

#### Rapid Literature Reviews

Two separate rapid literature reviews were conducted, one for CTD and one for GAMT, following the Cochrane rapid review methodology (Supplemental 2, 3).^47^ The protocols for the CTD and GAMT reviews were published on PROSPERO^48^ and Open Science Framework (OSF)^49^, respectively. Embase (via OVID) and MEDLINE (via OVID) were searched for relevant articles. Article screening, carried out on Covidence^50^, involved two reviewers (ZNM, JL) who independently screened the title, abstract, and full-text of each article. Articles not meeting the inclusion criteria (Supplemental 4) were excluded. Conflicts that could not be resolved between the reviewers were discussed with a third reviewer (SS-I). Data relating to outcomes, bibliographic information, and population details were extracted by one reviewer and verified by another.

#### CreatineInfo Patient Registry & Natural History Study

Additional outcomes were identified from the CreatineInfo Patient Registry and Natural History Study.^51^ CreatineInfo is a Cerebral Creatine Deficiency Syndrome (CCDS) patient- and caregiver-reported registry and natural history study created by the Association for Creatine Deficiencies (ACD), hosted by the National Organization for Rare Disorders (NORD). Outcomes were gathered from de-identified, aggregate data shared from CreatineInfo’s previously conducted Patient Meaningful Outcomes (PMO) survey which asked participants to identify and rate the outcomes that were most important to them.^52^

#### Focus Groups

In-person and virtual semi-structured focus groups were conducted with caregivers of individuals with CTD and GAMT to identify outcomes that were important to them. In-person focus groups took place at ACD’s annual symposium while virtual focus groups were conducted separately. Participants were recruited through their affiliation with the “Creatine Deficiency Support Group’’ and/or the “ACD Family Network”. One interpreter was present in one of the in-person focus groups.

Focus group participants were grouped according to proband diagnosis (CTD and GAMT) and age (0-8 years old and 9+ years old), when possible, for a total of five focus groups (three in-person and two virtual). Each focus group included three project team members: a facilitator who guided discussion and ensured equitable speaking time for each participant, an observer who served as quality control to the process and identified any bias that was introduced by the project team, and a notetaker. Participants discussed a series of broad questions designed to elicit their views on outcomes that would be important to them if improved with a treatment or therapy, based on their lived experiences (Supplemental 5).

The focus groups were audio-recorded and transcripts were generated using Otter.ai. The transcripts were cross-referenced with the audio recordings, all identifying information was removed, and focus group outcomes were extracted from the transcripts. These outcomes were added to the list of candidate outcomes from the rapid literature reviews and CreatineInfo. The list was then pruned by the project team to remove duplicate and non-disease-specific outcomes, with feedback from the PAReNts project caregivers and health professionals. A clear definition was created for the remaining outcomes in consultation with the PAReNts project caregivers and health professionals.

### Delphi Surveys

The goal of the Delphi survey phase was to systematically reduce the list of candidate outcomes by asking the CCDS community to rate and rank the importance of candidate outcomes.^53^ We conducted three Delphi survey rounds to collect data on which outcomes are most important to participants. We used the findings to move toward consensus on the list of candidate outcomes for our final COS.

#### Delphi Survey Development

Three sequential rounds of Delphi surveys (“Round 1”, “Round 2”, and “Round 3”) were created to evaluate the importance of candidate outcomes. Data were collected and managed using REDCap (Research Electronic Data Capture) hosted at Vanderbilt University.^54–55^ In all Rounds, participants were asked, for each outcome, “How important do you think it is for research studies to measure [outcome]” on a scale of 1-9, with 1-3 being of “limited importance”, 4-6 being “important but not critical”, and 7-9 being “most important”. Open text boxes were available in Rounds 1 and 2 for additional feedback. In Rounds 2 and 3, each outcome was accompanied by graphical distributions of ratings from the previous round by “Patients and Caregivers” and “Health Professionals”, and returning participants were provided with their own previous rating.

This process facilitated consensus on the outcomes, which is a key element of the Delphi methodology.^56^ In Round 3, participants also ranked their top 10 most important outcomes. Each survey included two catch trials that participants had to answer correctly for their entry to be included in the analysis. The surveys used lay language which was reviewed and improved through PAReNts review for clarity. This facilitated comprehension by all survey participants. Surveys were available in English, French, and Spanish, with translations completed by CryaCom International Inc. translation services and reviewed by native speakers.

#### Delphi Eligibility and Recruitment

Adult patients (18 years and older) who were able to express themselves and parents/caregivers of patients with CTD or GAMT were eligible to participate (“Patients and Caregivers”). Professionals with experience working with individuals with CTD and/or GAMT were also eligible to participate (“Health Professionals”). This included clinicians, researchers, laboratory scientists, health policy advisors, and additional professionals and therapists (e.g., speech therapists, psychologists, teachers). Round 1 was open to all eligible participants. Round 2 was open to Round 1 participants and new eligible participants. Round 3 was open exclusively to Round 1 and Round 2 participants.

Participants were recruited through direct email, newsletters, meetings, webinars, and social media. Recruitment materials were available in English, French, and Spanish. All recruitment materials for patients and caregivers were reviewed in advance by the PAReNts project caregivers. Active recruitment varied by survey round but lasted between 4-8 weeks per round. Participants received weekly email reminders to finish their surveys. Surveys were accessed via a publicly available link (Round 1) or by personalized survey links (Rounds 2 and 3).

#### Delphi Survey Analysis

At the completion of each survey round, the data were analyzed to determine which candidate outcomes were most important among stakeholders. Those outcomes that garnered the most support by reaching stakeholder consensus advanced to the next round. Stakeholder consensus was defined by the outcome inclusion criteria in each round, with the criteria becoming more rigorous in subsequent rounds. Inclusion criteria specifics are found in Figure 2. Criteria were applied first to “Patients and Caregivers” and “Health Professionals”; if the criterion was met for either of these groups, the outcome advanced. If an outcome did not meet the inclusion criterion for at least one of these two stakeholder groups, the subgroups “CTD Patients and Caregivers” and “GAMT Patients and Caregivers” were analyzed separately. An outcome had to meet the criterion for one of these subgroups to advance. In Round 3, inclusion criteria were applied in a stepwise manner (Figure 2; i.e., 3-1, 3-2, 3-3) in which an outcome had to meet all three criteria to advance. Upon completion of Round 3, the remaining list of candidate outcomes were further refined by combining similar outcomes and removing non-disease specific outcomes.

**Figure 2.**
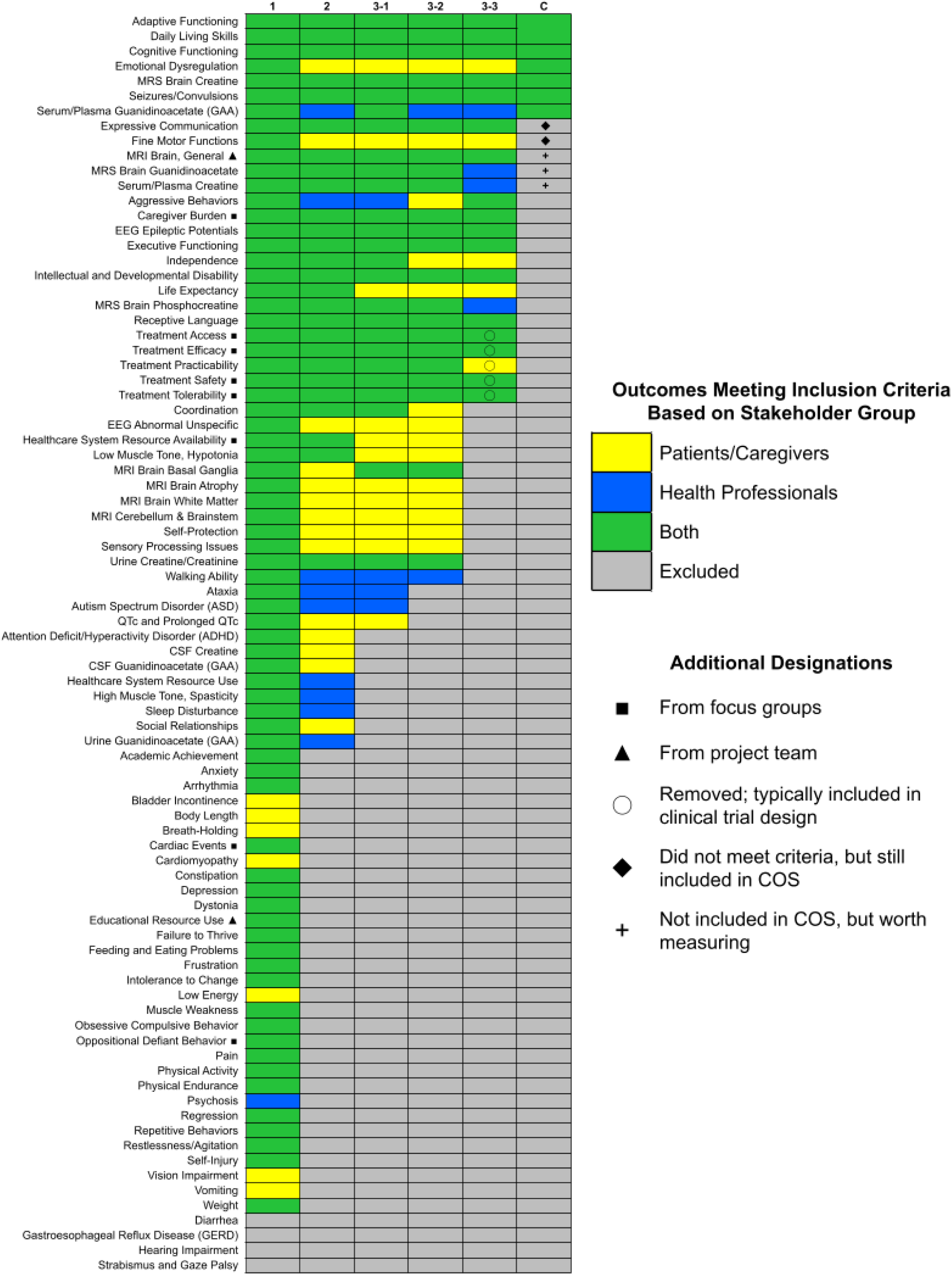
Outcomes remaining after each Delphi survey round and the consensus workshop. Column headers reflect the Delphi round and each set of inclusion criteria. The inclusion criteria for each round were as follows: Delphi 1, rated ≥3 by ≥70% of any stakeholder group; Delphi 2, rated ≥7 by ≥70% of any stakeholder group; Delphi 3-1, rated ≥7 by ≥70% of any stakeholder group; Delphi 3-2, mean rating ≥7 for any stakeholder group; Delphi 3-3, ranked in top 10 by ≥15% of any stakeholder group; Consensus (C), ≥50% of workshop participants voted outcome as “1-Definitely In”. Stakeholders meeting the inclusion criteria for each outcome are identified by color: patients/caregivers (yellow), health professionals (blue), both patients/caregivers and health professionals (green), outcome did not meet the criteria and was excluded (gray). Some outcomes were combined and/or changed throughout the process: “Developmental Delay” and “Intellectual Disability” were combined into “Intellectual and Developmental Disability”, “Adaptive Functioning” and “Daily Living Skills” were combined into “Adaptive Functioning”, and “Expressive Language” was changed to “Expressive Communication”. Outcomes marked with ◼ came exclusively from the focus groups. Outcomes marked with ▴ were introduced by the project team during the pruning phase. Outcomes marked with ○ were removed by the project team because they are typically already included in clinical trial designs. Outcomes marked with ◆ did not meet the consensus workshop inclusion criteria but were later included after unanimous agreement among participants. Outcomes marked with **+** are not included in the COS, but are worth measuring alongside the COS.

### Consensus Workshop

The goal of the consensus workshop was to prioritize the remaining candidate outcomes and reach consensus on a final list of 8-10 core outcomes that would become the COS for CTD and GAMT. The workshop was held in person at the University of Utah, Salt Lake City, Utah, USA; one member of the project team attended virtually. A group of 25 participants with a diverse range of perspectives attended the workshop to facilitate a holistic discussion of outcomes. Workshop participants consisted of CTD caregivers, GAMT caregivers, and health professionals/researchers including those who would use the COS in a clinical and research setting. The majority of workshop participants had completed at least one round of the Delphi survey and most of the caregivers were PAReNts cohort members. Two separate pre-workshop meetings were held for caregivers and health professionals to prepare participants for successful engagement in the workshop.

The workshop began with an overview of the COS development project and a review of Round 3 results. Utilizing an adapted Nominal Group Technique ^57^, each participant was then given one minute to provide their perspective on their three most important outcomes. Guided by a neutral moderator (BP), an open discussion unfolded among workshop participants. After thorough discussion of outcomes, each participant voted on the candidate outcomes via a Google Form. Caregivers voted once based on their lived experience with the disorder, while health professionals voted once for CTD and once for GAMT, based on their expertise. There were three response choices for each candidate outcome:

1) 1-Definitely In: outcome must be in the COS
2) 2-Maybe: outcome is a strong contender for the COS
3) 3-Definitely Out: outcome can be left out of the COS in favor of higher priority outcomes

Participants were asked to select a maximum of 5 outcomes as “1-Definitely In”. Once all participants had voted, the data were organized by disorder, analyzed using the consensus inclusion criteria, and shared with participants. Workshop participants discussed the voting results and eventually reached consensus on a final set of core outcomes.

## RESULTS

### Candidate Outcome Selection & Outcome Pruning

At the completion of the candidate outcome selection (i.e., literature review, focus groups, and CreatineInfo) 1148 outcomes were identified. After the outcome pruning stage (i.e., removal of duplicates and non-disease-specific outcomes, and feedback from caregivers and health professionals), 85 candidate outcomes remained.

Most of the 85 candidate outcomes were identified from multiple sources. Interestingly, 23 of the 85 outcomes were only found in the literature reviews, while eight were unique to the focus groups and not identified from any of the other sources. There were no unique outcomes identified only from CreatineInfo. Of the 85 outcomes, two emerged from discussions with the project team during the outcome pruning stage (Figure 2).

### Delphi Surveys

Collectively, 150 individuals participated in the Delphi surveys from 27 countries and six continents (Table 1). In Round 1, 120 participants rated 85 candidate outcomes. In Round 2, 81 remaining outcomes were rated by 111 participants, 32 of whom were new participants in Round 2 (i.e., retention rate from Round 1 = 71.2%). In Round 3, 84 participants rated the remaining 50 outcomes; 66 of the 84 participants (78.6%) had participated in Round 1, and 75 of the 84 participants (89.3%) had participated in Round 2. In each Delphi round, participants who did not answer at least 25% of the outcome survey questions or failed to correctly answer the catch trials were excluded from analysis. After Round 3, we further refined the candidate outcomes; similar outcomes were combined and five additional outcomes were removed because they are already typically included in clinical trial designs (Figure 2). No new outcomes were identified from comments shared by participants in open text boxes during Rounds 1 and 2. Detailed aggregate results from the Delphi survey, Supplemental 6.

**Table 1.**
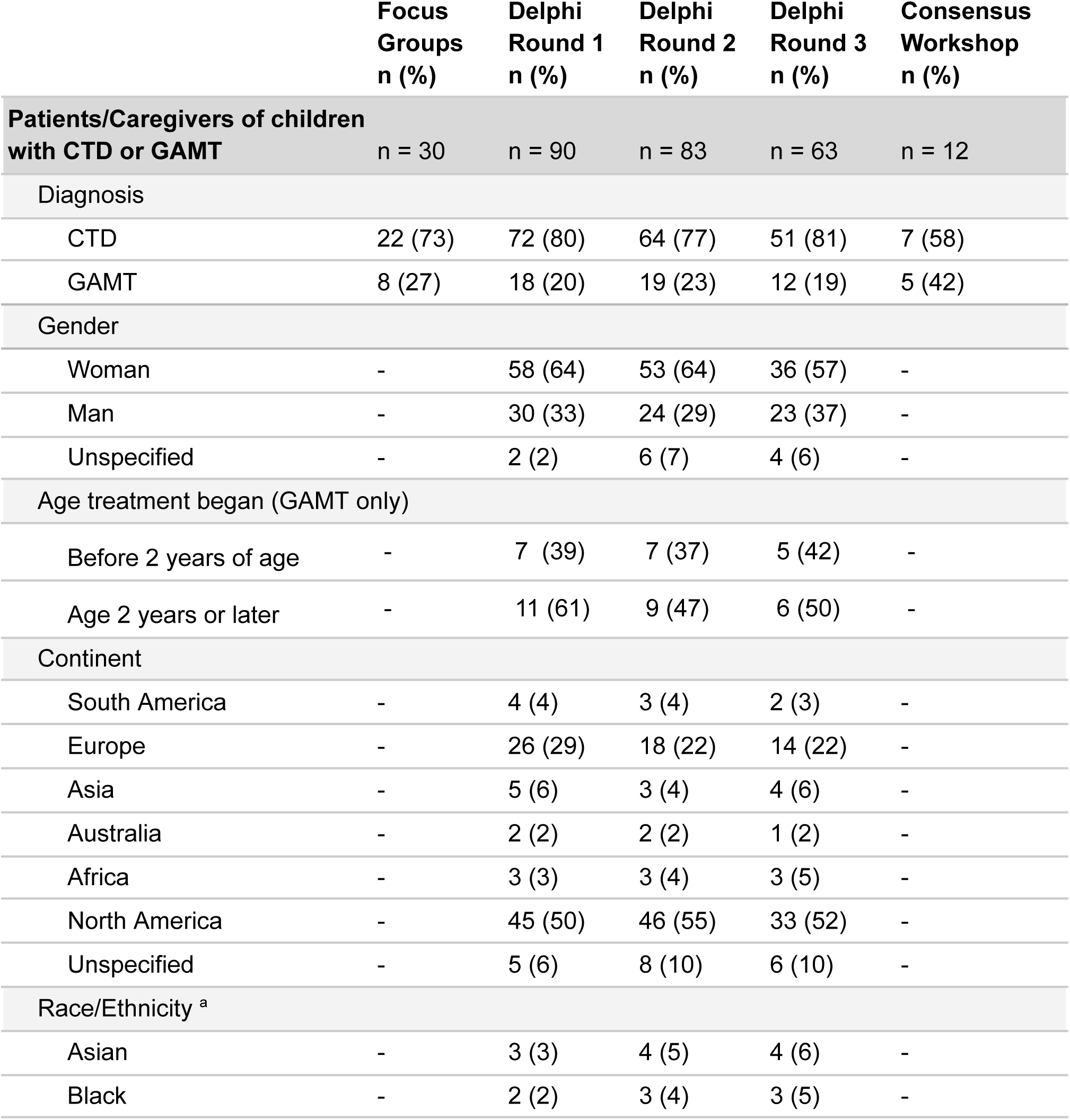
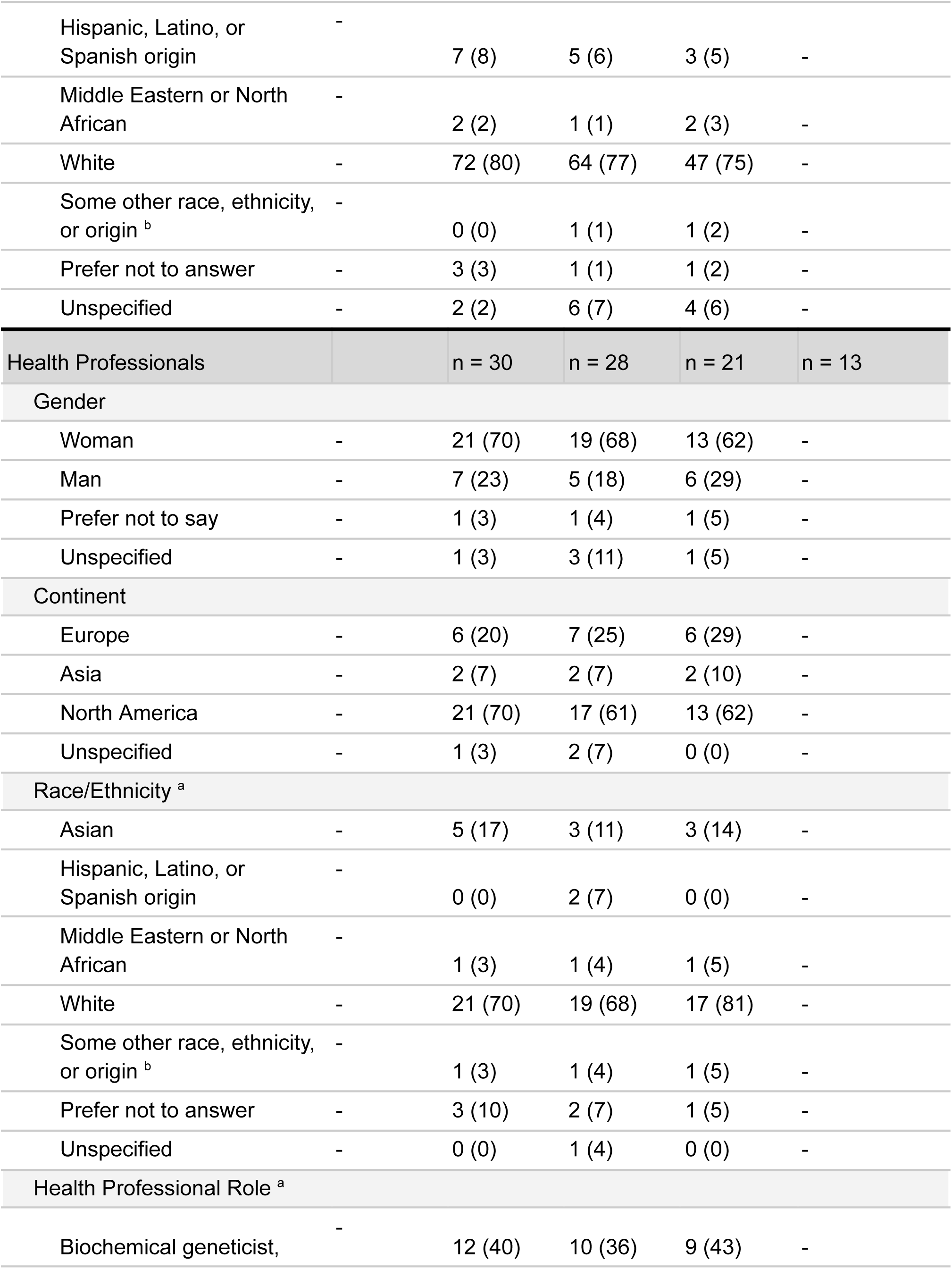
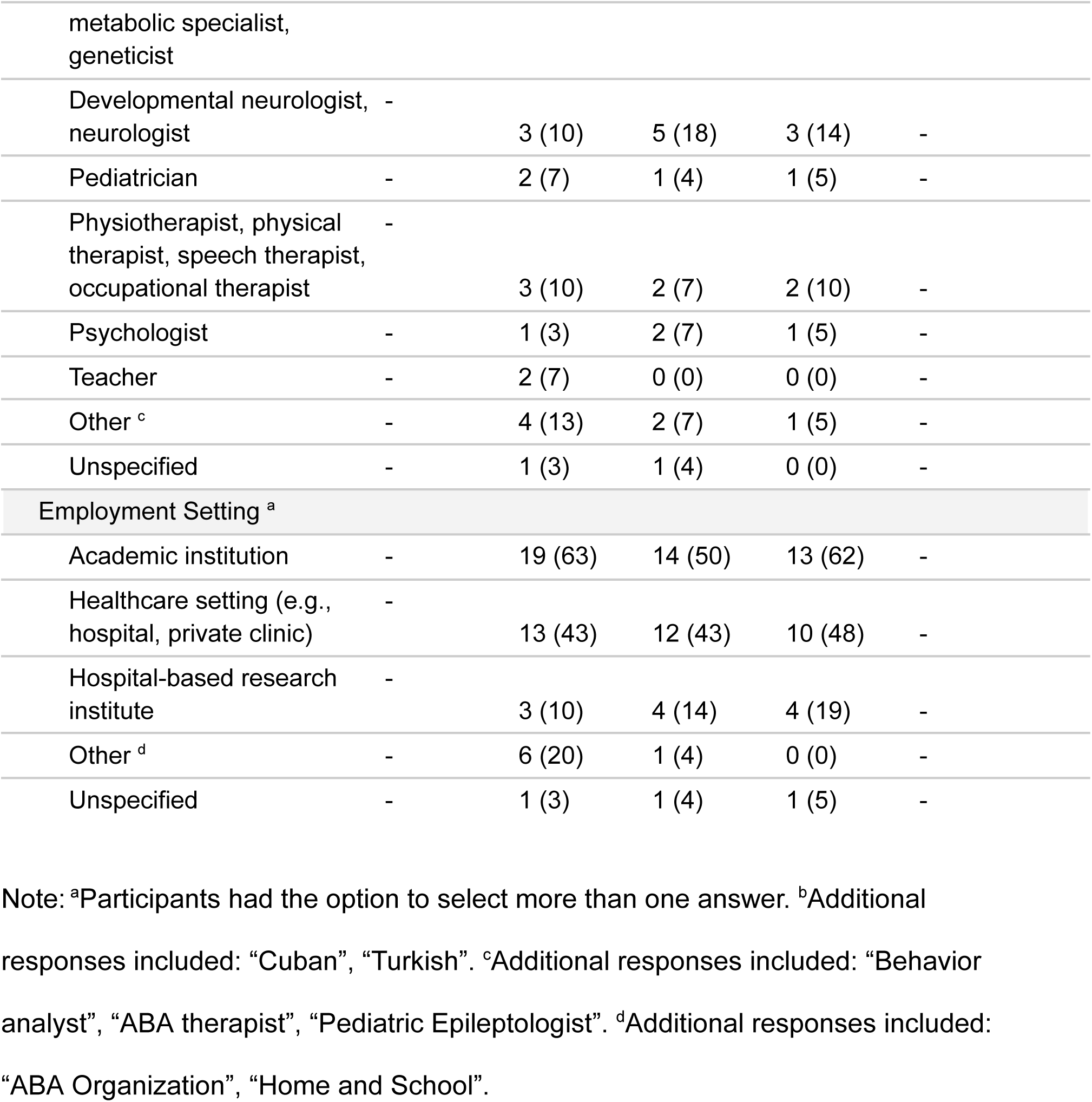
Demographics of focus groups, Delphi surveys, and consensus workshop participants.

### Consensus Workshop

Ahead of the consensus workshop, the 20 candidate outcomes were categorized into four core areas to highlight the comprehensive nature of our remaining candidate outcomes: “Life Impact”, “Growth and Development”, “Physiological/Clinical”, and “Death” (Table 2).^58–59^ Consensus workshop participants reviewed, discussed, and voted on the 20 candidate outcomes (Supplemental 7). Six outcomes were voted as “1-Definitely In” by more than 50% of workshop participants and were included in the COS: “Adaptive Functioning”, ”Cognitive Functioning”, “Emotional Dysregulation”, “MRS Brain Creatine”, “Seizure/Convulsions”, and “Serum/Plasma Guanidinoacetate” (for GAMT specifically). Two outcomes, “Expressive Communication” and “Fine Motor Functions”, did not meet the inclusion criteria but were included based on unanimous agreement during the group discussions. Thus, a total of eight outcomes (seven for CTD and GAMT, and one additional outcome for GAMT) were defined for the CTD and GAMT COS, with their definitions shown in Table 3. Three outcomes that were excluded from the COS were identified by workshop participants as worth tracking as they are often measured in parallel with the required COS: “MRI Brain General”, “MRS Brain Guanidinoacetate’’, and “Serum/Plasma Creatine” (Figure 2).

**Table 2.** Candidate outcomes discussed during the consensus workshop.

| Core area & definition | Outcomes discussed during consensus workshop (n=20) | Included in final COS? |
| --- | --- | --- |
| **Life Impact**<br>A general concept of well-being that refers to a combination of various life aspects [e.g., health, emotional, and social], as well as parents' perspectives about their involvement and observations | Aggressive Behaviors |  |
|  | Caregiver Burden |  |
|  | Emotional Dysregulation | yes |
|  | Seizure/Convulsions | yes |
| **Growth and Development**<br>process of change across a combination of physical, emotional, cognitive, social, and language areas over the course of childhood | Adaptive Functioning | yes |
|  | Cognitive Functioning | yes |
|  | Executive Functioning |  |
|  | Expressive Communication <sup>a</sup> | yes |
|  | Fine Motor Functions | yes |
|  | Independence |  |
|  | Intellectual & Developmental Disability |  |
|  | Receptive Language |  |
| **Physiological/Clinical**<br>observable and measurable abnormal changes and symptoms in the body as a result of CTD/GAMT | EEG Epileptic Potentials |  |
|  | MRS Brain Creatine | yes |
|  | MRI Brain General |  |
|  | MRS Brain Guanidinoacetate |  |
|  | MRS Brain Phosphocreatine |  |
|  | Serum/Plasma Creatine |  |
|  | Serum/Plasma Guanidinoacetate | yes |
| **Lifespan/Death**<br>the length/end of a person's life | Life Expectancy |  |

Note: <sup>a</sup>Outcome was originally called "Expressive Language" but was changed to
"Expressive Communication" during the consensus workshop.

**Table 3.**
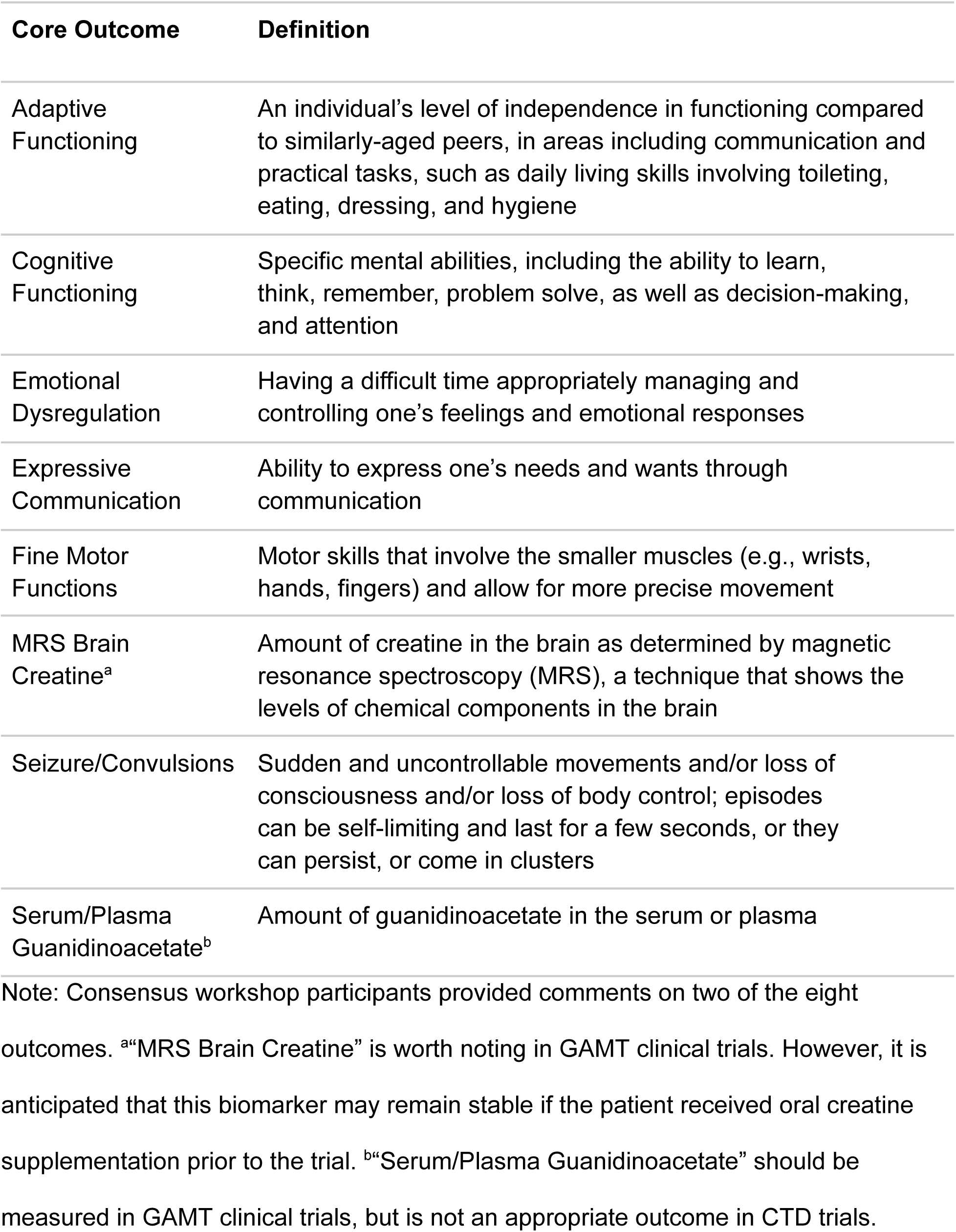
The final core outcome set (COS) for CTD and GAMT.

| **Core Outcome** | **Definition** |
| --- | --- |
| Adaptive Functioning | An individual's level of independence in functioning compared to similarly-aged peers, in areas including communication and practical tasks, such as daily living skills involving toileting, eating, dressing, and hygiene |
| Cognitive Functioning | Specific mental abilities, including the ability to learn, think, remember, problem solve, as well as decision-making, and attention |
| Emotional Dysregulation | Having a difficult time appropriately managing and controlling one's feelings and emotional responses |
| Expressive Communication | Ability to express one's needs and wants through communication |
| Fine Motor Functions | Motor skills that involve the smaller muscles (e.g., wrists, hands, fingers) and allow for more precise movement |
| MRS Brain Creatine<sup>a</sup> | Amount of creatine in the brain as determined by magnetic resonance spectroscopy (MRS), a technique that shows the levels of chemical components in the brain |
| Seizure/Convulsions | Sudden and uncontrollable movements and/or loss of consciousness and/or loss of body control; episodes can be self-limiting and last for a few seconds, or they can persist, or come in clusters |
| Serum/Plasma Guanidinoacetate<sup>b</sup> | Amount of guanidinoacetate in the serum or plasma |

Note: Consensus workshop participants provided comments on two of the eight outcomes. <sup>a</sup>“MRS Brain Creatine” is worth noting in GAMT clinical trials. However, it is anticipated that this biomarker may remain stable if the patient received oral creatine supplementation prior to the trial. <sup>b</sup>“Serum/Plasma Guanidinoacetate” should be measured in GAMT clinical trials, but is not an appropriate outcome in CTD trials.

During the consensus workshop discussion, participants removed the outcomes “Intellectual & Developmental Disability”, “Receptive Language”, “Independence”, and “Executive Functioning”. While “Intellectual & Developmental Disability” is a prominent diagnosis in the population, its components, “Cognitive Functioning” and “Adaptive Functioning”, were included in the COS and can be measured with metrics more sensitive to change. “Independence” is reflected in “Adaptive Functioning” while “Executive Functioning” is an aspect of “Cognitive Functioning”. “Expressive Language” was renamed “Expressive Communication” to include non-verbal forms of communication.

## DISCUSSION

Using a structured consensus process, we successfully identified a set of eight core outcomes that should be used in clinical trials and research studies for CTD and GAMT based on caregiver and health professional input. Following the recommended COMET framework, we identified candidate outcomes by performing literature reviews, focus groups, and Delphi surveys, and finalized the COS by holding an in-person consensus workshop.^41^ The final COS includes outcomes from three of the relevant recommended core areas, meeting the current recommendation that a COS be holistic to better represent the patient.^58–59^

Seven of the eight outcomes are applicable to both CTD and GAMT. “Serum/Plasma Guanidinoacetate” is a unique outcome exclusive to GAMT because elevated GAA levels are a hallmark of GAMT and reducing GAA is a standard objective of GAMT treatment. In contrast, GAA levels tend to be normal in individuals with CTD and are not expected to change with any treatment. Measurement of creatine with magnetic resonance spectroscopy (MRS), “MRS Brain Creatine”, is recommended for CTD clinical trials as cerebral creatine is markedly low in CTD and may increase with a successful treatment. This outcome is also recommended for monitoring in the case of GAMT; however this biomarker may be normalized in patients receiving oral creatine supplementation prior to a trial with a new therapy.

Workshop participants agreed that three outcomes eliminated from the core outcome set will often be measured in parallel with the required COS and are worth tracking. In the case that plasma is collected for “Serum/Plasma Guanidinoacetate”, tracking “Serum/Plasma Creatine” is encouraged in trials that aim to replenish GAMT activity and thus endogenous synthesis of creatine. Similarly, when measuring “MRS Brain Creatine”, also measuring “MRS Brain Guanidinoacetate’’ may provide valuable monitoring insights in GAMT trials. When magnetic resonance imaging (MRI) is performed in parallel with MRS, any abnormality in “MRI Brain General” in GAMT and CTD clinical trials is recommended to be noted. This is supported by the fact that certain abnormalities (e.g. abnormal signal intensity in basal ganglia) are reversible in patients with GAMT upon treatment with creatine and/or guanidinoacetate lowering therapies.^19,60^

### Caregiver involvement in developing a COS for two rare conditions

CTD and GAMT present with similar phenotypes and share the treatment goal of restoring creatine to the brain.^11^ Additionally, patients, caregivers, researchers, and clinicians actively partner together to advance research on these two conditions. Given their ultra-rare nature, both conditions benefit from mutual support within this collaborative community. We hypothesized that very few critical differences in prioritized outcomes would be identified between these two conditions, thereby driving our effort to develop a COS for CTD and GAMT simultaneously. Our COS supports this hypothesis, with seven of the core outcomes applicable to both conditions.

Though we utilized multi-stakeholder collaboration to develop a COS, we took deliberate measures to ensure that caregiver and patient voices remained distinct. Our caregiver focus groups resulted in eight candidate outcomes that were exclusively identified by caregivers, with one outcome (“Caregiver Burden”) receiving extensive discussion during the consensus workshop (see Figure 2). By conducting caregiver focus groups and analyzing Delphi responses separately for each diagnosis and stakeholder group, we created a COS that is informed by the experiences of CTD and GAMT patients and families. For example, the impacts of “Seizures/Convulsions”, a frequent characteristic of both CTD and GAMT^11^, were highlighted during the consensus workshop when one caregiver conveyed the effects on their family:

> *“Ninety-nine percent of my burden is my daughter’s seizures and her moods that come out of nowhere because she’s going to have a seizure. And that they ruin my family’s life."*

Similarly, many caregivers expressed concern about their child(ren)’s “Emotional Dysregulation”, an outcome that encompasses an array of emotional responses such as aggression and irritability. One caregiver expressed during the focus group:

> *“Last night, I had an iPad thrown at my face. He gets mad really quickly. He can’t seem to control his outbursts. [He’s] put his foot through a wall before.”*

Impaired “Expressive Communication” was another key concern of many caregivers. One parent shared their frustration when their child is unable to verbally communicate with others:

> *"I can see that he gets disappointed when he cannot get his message across to others. And it’s frustrating for me because I’m trying to understand him, but I can’t.”*

These real-world examples demonstrate how caregivers’ experiences and their related outcomes are captured in the COS. Patient and caregiver participation in our project was integral to developing a COS that is not only disease-specific but also patient-centered.

While consensus on outcomes was eventually reached, differences in outcome prioritization between caregivers and health professionals were identified during the Delphi survey and consensus workshop phases (Figure 2). For example, “Emotional Dysregulation” and “Fine Motor Skills” both met the inclusion criteria among caregivers, but not health professionals in Delphi Rounds 2 and 3. While “Emotional Dysregulation” met the consensus workshop inclusion criteria, “Fine Motor Skills” initially did not. After further discussion, workshop participants unanimously agreed to include both “Emotional Dysregulation” and “Fine Motor Skills” in the final COS. Conversely, “Serum/Plasma Guanidinoacetate (GAA)” met the inclusion criteria among health professionals, but not caregivers, in Delphi Rounds 2 and 3. During the consensus workshop discussions, participants reached a unanimous agreement to include it in the final COS. Additionally, “Caregiver Burden” was an outcome that received thoughtful debate among consensus workshop participants. Caregivers argued that improvements in patient-focused outcomes will naturally lead to reduced “Caregiver Burden”, making inclusion of this outcome unnecessary. Health professionals agreed to defer to caregivers and this outcome was not included in the COS.

Caregivers utilized their research engagement training to effectively collaborate with health professionals and advocate for their families during the consensus workshop, directly influencing the final COS. For instance, caregivers voiced their concern that “Expressive Language” and its original definition was too limited and exclusionary, failing to capture meaningful improvements in non-verbal forms of communication (e.g., sign language, communication devices). Participants collectively agreed to change this outcome to “Expressive Communication” and modify the definition to include non-verbal forms of communication. Throughout the COS development process, caregivers’ lived experiences complemented the health professionals’ clinical expertise, resulting in an inclusive and holistic COS for CTD and GAMT.

### COS Importance in CTD and GAMT clinical trials

Given the rarity of CTD and GAMT, it is essential that researchers use this multistakeholder-influenced COS in every clinical trial for these conditions as it effectively captures what is most important for these patients. This COS will become increasingly impactful as new treatments are developed, as the COS represents an opportunity to ensure that these therapies are evaluated based on meaningful outcomes. Additionally, using the COS contributes to reduced waste, standardizes comparison across studies, and promotes collaboration and data sharing. We expect our COS to increase the efficiency of research methods and clinical trial designs, and ultimately expedite the development of effective treatments for CTD and GAMT. Moreover, regulatory agencies, such as the U.S. FDA, can be reassured that patient perspectives are included in the proposed trial design when the COS is included. By including patient and caregiver perspectives to establish a COS for CTD and GAMT, the investigator burden of creating patient-centered trial design is greatly reduced.

### Recommendations for COS Development and Future Directions

This collaboration marks the first COS developed for CTD and GAMT, and is one of only a few efforts to create a COS for inborn errors of metabolism (IEM). Our approach shares many similarities with other COS development projects for IEM, including mucopolysaccharidoses (MPS)^61–62^, phenylketonuria (PKU) and medium chain acyl coA dehydrogenase deficiency (MCAD)^63^, as they were also guided by the COMET framework. Our project is unique compared to other COS development projects, as it incorporated all of the following design elements: 1) caregiver and health professional participation, 2) focus groups, 3) literature reviews, 4) patient registry data, 5) Delphi surveys, and 6) consensus workshop. Other unique features instrumental to our consensus process included progressively more rigorous Delphi inclusion criteria, approximately equal numbers of caregiver and health professionals in the consensus workshop, and our workshop voting process.

We hope that other disease populations, especially other rare diseases, will benefit from the strengths of our COS development project and our recommendations on potential areas for improvement as they develop their own COS. One of the unique components of this project was the active engagement of caregivers in the entire process, as part of our larger PAReNts Project. Caregivers received training and education in COS and clinical trial development, learned how to become better advocates by sharing their personal stories, and contributed to the focus groups, Delphi surveys, and consensus workshops. Health professionals commented on how knowledgeable caregivers were during the workshop discussions:

> *”Their ability to synthesize and comment on the scientific issues was amazing.”*

Based on our experience, we propose the use of adaptable inclusion criteria for retainment of proposed outcomes between Delphi survey rounds. Many patients and caregivers find it challenging to deprioritize outcomes^41^, which may result in limited variability in the scoring of outcomes using less rigorous *a priori* criteria. After thoughtful consideration, we implemented more rigorous *post hoc* criteria in Delphi Rounds 2 and 3 to filter the total number of outcomes. Reducing outcomes prior to survey distribution decreases the burden on participants and the attrition rate between rounds.^41,64^

We reflect that the pre-workshop training and the in-person format of our consensus workshop, with approximately equal numbers of caregivers and health professionals in attendance, helped us reach consensus on the COS more efficiently. Additionally, the alternating placement of stakeholders throughout the consensus workshop room facilitated open conversation with all participants and encouraged them to make new connections.

Future COS development projects should prepare for potential challenges. In the case of stakeholder participation, it is important to target recruitment of stakeholders who are from traditionally underrepresented groups to ensure their participation and minimize bias. For example, GAMT is rarer than CTD; therefore, we employed various recruitment strategies to ensure a more representative participant pool between the two conditions, still not reaching equal representation. We recommend inclusion of health policy advisors, as we believe this would have provided us with valuable insights on clinical trials and the drug development process during this project. It is important to consider in advance how to accommodate the communication needs of participants.

Our materials only included English, French, and Spanish translations; additional languages would have broadened participation. It is important to consider translating recruitment documents, project materials, and the Delphi survey into multiple languages, and providing interpretation services or closed captioning during focus groups and the consensus workshop.

While the outcomes established in the COS serve as a foundation, we acknowledge that the measurement of additional outcomes may be beneficial depending on the type of therapeutic intervention, characteristics of the target patient population, and other variables. For instance, phenotypes of males and females with CTD may vary significantly and warrant the inclusion of additional outcomes to capture these differences. An N-of-1 trial may benefit from additional customized outcomes as well.

Understanding how to measure the COS is an important next step. Outcomes may be measured using different tools. Not all tools may be effective in a particular population, especially among neurodevelopmentally impacted patient populations like CTD and GAMT. Development of patient-centered considerations for selecting outcome measurement tools appropriate to this patient community is a logical next step and will serve as a valuable companion to our COS.^41^

In order to design appropriate and relevant studies and clinical trials for rare diseases, it is important to have a group of patient-centered core outcomes that are consistently measured to enable comparison across studies and facilitate therapeutic development. Here, we share the first COS for CTD and GAMT. Throughout the development of this COS, patient and caregiver perspectives were considered through their engagement in focus groups, Delphi surveys, and the consensus workshop. We especially benefited from the caregivers who partnered with us to co-design the recruitment materials, outcome definitions, Delphi surveys, and attended the consensus workshop. This COS is a step towards designing more appropriate clinical trials and accelerating the development of effective interventions for CTD and GAMT.

## ACKNOWLEDGMENTS

We thank our PaReNts Project caregiver collaborators, who contributed a wealth of knowledge and perspectives through their lived experiences to all aspects of this project: Irene Adjetey, Regina Bogar, Rachel & Mac Cafferty, Carole Chehowah, Daniel Coller, Reza Fathi, Celeste Graham, Vicky Hall, Mikelle Law, Gustavo Mattos, Gina Nichols, Haley Prescher, Beth Robinson, Anthony Tedesco, Beth Tiller, Kim Tuminello, Nathan Vandenberg, Lacy Vandivort, Celine Wheaton, and Kayla & Sidney Williams. We also thank everyone who participated in the Delphi surveys. Thank you to Carole Chehowah and Laura Duque Lasio for their assistance in translating Delphi survey materials. Thank you to Alison Howie and Sangeetha Iyer for their support in developing this project. Thank you to Cristan Farmer and Andrew Marshall for their help in building this manuscript.

## AUTHOR CONTRIBUTIONS

AT, BP, MS, SB, TS, SS-I, and HW conceptualized the project; ZNM, EKR, AT, BP, MS, CG, BHT, SS-I, and HW designed the project; MS, CG, BHT, and HW led the caregiver engagement strategy as patient/caregiver partners; ZNM, EKR, JL, SS-I, and HW acquired the data; ZNM, EKR, AT, BP, MS, CG, BHT, SAB, DAB, RB, JB, RC, DPC, TJDG, VH, GSL, NL, SM-A, JSM, MP, GSS, AS, CPW, KFW, SPY, SS-I, and HW analyzed and interpreted the data; ZNM, EKR, AT, SS-I, and HW drafted the manuscript; BP, MS, CG, BHT, SAB, DAB, RB, JB, RC, DPC, TJDG, VH, GSL, NL, SM-A, JSM, MP, GSS, AS, CPW, KFW, and SPY critically revised the manuscript. All authors approve the final manuscript.

## DATA AVAILABILITY STATEMENT

Raw data are not publicly available due to privacy and/or ethical restrictions.

Aggregate data that supports the findings of this study are available in the supplementary materials of this article. Additional data may be made available on request from the corresponding author. Data ownership is maintained by ACD.

## ETHICS STATEMENT

All focus groups and Delphi surveys received institutional review board (IRB) approval from the North Star Review Board (protocols NB200065, NB300094; https://learningirb.org/).

## PATIENT INFORMED CONSENT

Informed consent was obtained from all focus group and Delphi survey participants prior to participation (IRB protocols NB200065, NB300094).

