## Supplemental Materials for "Establishing a Core Outcome Set for Creatine Transporter Deficiency and Guanidinoacetate Methyltransferase Deficiency"

**SUPPLEMENTARY MATERIAL**

Supplemental 1. Core Outcome Set-STandards for Reporting: The COS-STAR Statement Checklist

| **Section/Topic** | **Item No.** | **Checklist Item** | **Reported On Page Number** |
|---|---|---|---|
| Title/Abstract | | | |
| Title | 1a | Identify in the title that the paper reports the development of a COS | 1 |
| Abstract | 1b | Provide a structured summary | 2-3 |
| Introduction | | | |
| Background and Objectives | 2a | Describe the background and explain the rationale for developing the COS. | 4-5 |
| | 2b | Describe the specific objectives with reference to developing a COS. | 4-5 |
| Scope | 3a | Describe the health condition(s) and population(s) covered by the COS. | 4-5 |
| | 3b | Describe the intervention(s) covered by the COS. | 4-5 |
| | 3c | Describe the setting(s) in which the COS is to be applied. | 4-5 |
| Methods | | | |
| Protocol/Registry Entry | 4 | Indicate where the COS development protocol can be accessed, if available, and/or the study registration details. | 6 & 41 |
| Participants | 5 | Describe the rationale for stakeholder groups involved in the COS development process, eligibility criteria for participants from each group, and a description of how the individuals involved were identified. | 6, 8, 10- |
| Information Sources | 6a | Describe the information sources used to identify an initial list of outcomes. | 7-8, 13, 32, Figure 1 |
| | 6b | Describe how outcomes were dropped/combined, with reasons (if applicable). | 9-11, 13-14, 32, Figure 1, 33-34, Figure 2 |
| Consensus Process | 7 | Describe how the consensus process was undertaken. | 11-12, 14 |
| Outcome Scoring | 8 | Describe how outcomes were scored and how scores were summarised. | 9-11 |

| | | | |
|---|---|---|---|
| Consensus Definition | 9a | Describe the consensus definition. | 11 |
| | 9b | Describe the procedure for determining how outcomes were included or excluded from consideration during the consensus process. | 11-12 |
| Ethics and Consent | 10 | Provide a statement regarding the ethics and consent issues for the study. | 25-26 |
| Results | | | |
| Protocol Deviations | 11 | Describe any changes from the protocol (if applicable), with reasons, and describe what impact these changes have on the results. | 22 |
| Participants | 12 | Present data on the number and relevant characteristics of the people involved at all stages of COS development. | 27-29, Table 1 |
| Outcomes | 13a | List all outcomes considered at the start of the consensus process. – table 2 | 30, Table 2 |
| | 13b | Describe any new outcomes introduced and any outcomes dropped, with reasons, during the consensus process. | 16-17, 19-20, 30, Table 2 |
| COS | 14 | List the outcomes in the final COS. | 14, 31, Table 3 |
| Discussion | | | |
| Limitations | 15 | Discuss any limitations in the COS development process. | 22-23 |
| Conclusions | 16 | Provide an interpretation of the final COS in the context of other evidence, and implications for future research. | 23-24 |
| Other Information | | | |
| Funding | 17 | Describe sources of funding/role of funders. | 25 |
| Conflicts of Interest | 18 | Describe any conflicts of interest within the study team and how these were managed. | 3 |

Note: This table was sourced from the paper published by Kirkham et al. which provides guidance for reporting of all COS studies.[45]

Supplemental 2. Rapid literature review outcome extraction sheet

| Literature Review Outcome Extraction Sheet | | | | | | | | | | | | | | | | | | | | | | |
|---|---|---|---|---|---|---|---|---|---|---|---|---|---|---|---|---|---|---|---|---|---|---|
| BIBLIOGRAPHICAL INFORMATION | | | | | | | | | POPULATION DETAILS | | | | | | INTERVENTION /EXPOSURE DETAILS | OUTCOME DETAILS (add a row for each additional outcome) | | | | Extracted by | Verified by | Verification notes |
| Covidence ID | Title | Year | First Author (Last Name) | Journal | Country of Study | Study Design | Single/multi centre, single/multi country | Time period | Sample Size | Age - Diagnosed | Age - Symptoms Occured | Disease Severity | Family History/Affected Siblings | Ethnicity | Describe the intervention/exposure | Outcome details | Describe scale/tool used to measure outcome | Was the scale/tool validated? | Reference for validation | | | |
| Note: authors of the included papers were not contacted for missing or unclear data. | | | | | | | | | | | | | | | | | | | | | | |

Note: This supplemental file includes the extraction sheet used to record outcomes identified through our CTD and GAMT literature reviews

Supplemental 3. Rapid literature review search strategies

| **CTD Search Strategy** | |
|---|---|
| Set | Search Statement |
| 1 | *Mental Retardation, X-Linked/ or *Creatine/ |
| 2 | (CRTD or creatine transporter* deficienc* or creatine transporter* activit*).mp. |
| 3 | SLC6A8*.mp. |
| 4 | 1 or 2 or 3 |
| 5 | patient reported outcome measures/ |
| 6 | Outcome Assessment, Health Care/ |
| 7 | Treatment Outcome/ |
| 8 | ((patient* or core) adj3 (outcome* or measure*)).tw,kf. |
| 9 | (outcome* adj3 (assessment* or treatment* or therapy*)).tw,kf. |
| 10 | (prom or proms).tw,kf. |
| 11 | or/5-10 |
| 12 | Intellectual Disability/ or Developmental Disabilities/ or Developmental Delay/ |
| 13 | (intellect* disabilit* or learn* disabilit* or development* disabilit* or development* delay* or development* regress*).tw,kf. |
| 14 | Epilepsy/ or Seizures/ or Seizures, Febrile/ |
| 15 | (epilep* or seizure*).tw,kf. |
| 16 | (speech adj3 (delay* or impediment*)).tw,kf. |
| 17 | Movement Disorders/ or Muscle Hypotonia/ or Muscle Weakness/ or "Activities of Daily Living"/ |
| 18 | (movement disorder* or muscle hypotonia* or muscle weakness* or activit* of daily living*).tw,kf. |
| 19 | (dystonia* or spasticit* or parkinsonism* or rigidit* or ataxia* or chorea* or asteatos*).tw,kf. |
| 20 | dementia*.tw,kf. |
| 21 | Autism/ or ADHD/ or ASD/ |
| 22 | (autis* spectrum disorder* or autism or attention* deficit* hyperactivity* disorder*).tw,kf. |
| 23 | Problem Behavior/ |
| 24 | (problem adj3 (behaviour* or behavior*)).tw,kf. |
| 25 | Mental Disorders/ or Anxiety/ or Depression/ or OCD/ or Schizophrenia/ |
| 26 | (mental disorder* or mental illness* or anxiety or depression or obsessive compulsive disorder* or OCD or schizophrenia).tw,kf. |

| | |
|---|---|
| 27 | Insomnia/ or Apnea/ or Parasomnia/ or Nocturnal Enuresis/ |
| 28 | (insomnia* or apnea* or parasomnia* or nocturnal enuresis*).tw,kf. |
| 29 | Feeding Behavior/ |
| 30 | (feeding adj3 (behaviour* or behavior*)).tw,kf. |
| 31 | Pain Measurement/ or Pain Management/ or Pain/ |
| 32 | (pain* or pain tolerance or hyperacusis or sensory processing disorder or food avers* or hypersensitiv* or hyperalgesia).mp. [mp=title, abstract, heading word, drug trade name, original title, device manufacturer, drug manufacturer, device trade name, keyword heading word, floating subheading word, candidate term word] |
| 33 | Vision/ or Hearing/ or Sensory/ or Somatosensory Disorders/ |
| 34 | (vision or hearing or sensory).tw,kf. |
| 35 | Osteoblasts/ or Osteoporosis/ or Bone Diseases/ |
| 36 | (bone density or osteopenia or osteoporosis or osteoblast* or bone disease*).tw,kf. |
| 37 | Gastrointestinal Diseases/ |
| 38 | (gastrointestinal disease* or gastrointestinal symptom* or constipation or acid reflux or GERD or gastroesophageal reflux disease).tw,kf. |
| 39 | (weight or height).tw,kf. |
| 40 | "Quality of Life"/ |
| 41 | (quality of life or life quality).tw,kf. |
| 42 | exp Communication Disorders/ |
| 43 | communicat*.tw,kf. or expressive language delay*.mp. or speech delay*.mp. or delay in speech.mp. or nonverbal communicat*.mp. or nonverbal.mp. or social communicat*.mp. or emotional communicat*.mp. or sign language.mp. or assisted communicat*.mp. or alternative communicat*.mp. or augumentative communicat*.mp. [mp=title, abstract, heading word, drug trade name, original title, device manufacturer, drug manufacturer, device trade name, keyword heading word, floating subheading word, candidate term word] |
| 44 | exp Sleep Wake Disorders/ |
| 45 | (sleep wake disorder* or sleep disorder* or sleep abnormalit* or insomnia* or parasomnia* or nocturnal enuresis or sleep disordered breathing or sleep disordered grinding or sleep apnea or restless sleep* or sleep restlessness or sleep restriction or hypersomnia).tw,kf. |
| 46 | or/12-45 |
| 47 | 11 or 46 |
| 48 | Creatine/ or phosphocreatine/ or exp Creatine Kinase/ or Biomarkers/ |
| 49 | (Creatine or phosphocreatine or creatine phosphate or creatine kinase or creatine phosphokinase or Brain creatine or urine creatine or guanidinoacetate or CK or CPK).tw,kf. |

| | |
|---|---|
| 50 | 48 or 49 |
| 51 | 4 and 47 |
| 52 | 4 and 47 and 50 |
| **GAMT Search Strategy** | |
| Set | Search Statement |
| 1 | Guanidinoacetate N-Methyltransferase/ |
| 2 | (guanidinoacetate methyltransferase deficien* or GAMT deficien* or cerebral creatine deficiency).mp. |
| 3 | (creatine adj3 (deficien* or disorder*)).mp. |
| 4 | 1 or 2 or 3 |
| 5 | intellectual disability/ or developmental disability/ or developmental delay/ or developmental disabilities/ |
| 6 | ((intellectual* or neurologic* or mental* or cerebral* or neurodevelopment* or neuro-development* or development* or learn* or cognitive or cognition) adj3 (impair* or dysfunction* or delay* or disabilit* or disorder* or regress* or deficit*)).mp. |
| 7 | (global developmental delay* or GDD).mp. |
| 8 | epilepsy/ or seizures/ or seizure, febrile/ |
| 9 | (epileps* or epileptic encephalopath* or seizure*).mp. |
| 10 | Dementia/ |
| 11 | dementia*.mp. |
| 12 | Mental Disorders/ or Anxiety/ or Depression/ or OCD/ or Schizophrenia/ |
| 13 | (mental disorder* or mental illness* or anxiety or depression or obsessive compulsive disorder* or OCD or schizophrenia or depressed or anxious or compulsive behavior* or compulsive behaviour*).mp. |
| 14 | Autism/ or Autism Spectrum Disorder/ or ASD/ or ADHD/ |
| 15 | (autism* or attention deficit* hyperactiv* disorder* or ADHD or ASD).mp. |
| 16 | Problem Behavior/ |
| 17 | ((aggression or aggressive or problem*) adj3 (behaviour* or behavior*)).mp. |
| 18 | ((abnormal or self-mutilat* or self mutilat* or selfmutilat* or auto-mutilat* or auto mutilat* or automutilat* or self-injur* or self injur*) adj3 (behavior* or behaviour*)).mp. |
| 19 | Communication Disorders/ |
| 20 | (communicat* or expressive language delay* or speech delay* or speech impediment* or delay in speech or nonverbal or non-verbal or social communicat* or emotional communicat* or sign language or ASL or assist* communicat* or alternat* communicat* or augumentat* communicat*).mp. |
| 21 | ((Absent or slurred) adj3 (speech or language)).mp. |

| | |
|---|---|
| 22 | ((speech or language) adj3 (impediment* or delay* or impairment*)).mp. |
| 23 | (no language or no speech).mp. |
| 24 | Feeding Behavior/ |
| 25 | ((feed* or eat*) adj3 (behaviour* or behavior*)).mp. |
| 26 | Movement Disorders/ or Muscle Hypotonia/ or Muscle Weakness/ or "Activities of Daily Living"/ |
| 27 | ((motor or movement*) adj3 (impair* or dysfunction* or delay* or disabilit* or disorder* or regress* or deficit*)).mp. |
| 28 | (stiff posture* or abnormal posture*).mp. |
| 29 | (dysarthri* or hyperreflexia* or hyper-reflexia* or spasticit* or spastic or ataxia* or ataxic or dystoni* or hypotoni* or hypo-toni* or Parkinson* or Pes Cavus or choreoathetosis or choreo-athetosis).mp. |
| 30 | (myopath* or neuropath*).mp. |
| 31 | (delay* adj3 (sit* or stand* or walk*)).mp. |
| 32 | ((muscle* or muscular) adj3 (reduc* or decrease* or atroph* or weakness or hypotoni* or hypo-toni*or dystoni* or dystroph*)).mp. |
| 33 | (activit* of daily living* or ADL*).mp. |
| 34 | arthritis/ |
| 35 | (arthrit* or osteoarthrit* or osteo-arthrit*).mp. |
| 36 | Osteoblasts/ or Osteoporosis/ or Bone Diseases/ |
| 37 | (bone fracture* or bone deformit* or osteoporosis or osteopenia* or bone densit* or osteoblast* or bone disease*).mp. |
| 38 | Sleep Wake Disorders/ or Insomnia/ or Apnea/ or Parasomnia/ or Nocturnal Enuresis/ |
| 39 | (sleep wake disorder* or sleep-wake disorder* or sleep disorder* or sleep abnormalit* or insomnia* or parasomnia* or nocturnal enuresis or sleep disordered breathing or sleep disordered grinding or sleep apnea or restless sleep* or sleep restlessness or sleep restriction* or hypersomnia* or apnea).mp. |
| 40 | (extrapyramidal syndrome or extrapyramidal dysfunction or involuntary movement* or drool* or salivat*).mp. |
| 41 | chorea*.mp. |
| 42 | Pain Measurement/ or Pain Management/ or Pain/ |
| 43 | (pain* or pain toleran* or hyperacusis or sensory processing disorder* or food avers* or hypersensitiv* or hyperalgesi*).mp. |
| 44 | Vision/ or Hearing/ or Sensory/ or Somatosensory Disorders/ |
| 45 | (vision or hearing or sensory or somatosensor* or somato-sensor*).mp. |
| 46 | Gastrointestinal Diseases/ |

| | |
|---|---|
| 47 | (gastrointestinal disease* or gastro-intestinal disease* or gastrointestinal symptom* or gastro-intestinal symptom* or constipat* or diarrhea* or acid reflux* or GERD or gastroesophageal reflux diseas* or gastro-esophageal reflux disease* or growth retard* or failure to thrive or short stature).mp. |
| 48 | body height/ or body weight/ |
| 49 | (body weight or body height or head circumference*).mp. |
| 50 | "Quality of Life"/ |
| 51 | (quality of life or quality-of-life or life quality or life-quality or QOL).mp. |
| 52 | or/5-51 |
| 53 | Biomarkers/ |
| 54 | (Biomarker* or biochemical marker* or test* or assess* or GAA or guanidinoacetate or creatinine or Glycine or MRI or MRS or magnetic resonance imaging or magnetic resonance spectroscopy or elevated GAA* or elevated level* of GAA or serum creatine or GAA level* or biochemical assay*).mp. |
| 55 | Creatine/ |
| 56 | (creatine adj3 (supplement* or excret* or deplet*)).mp. |
| 57 | patient reported outcome measures/ or Outcome Assessment, Health Care/ or Treatment Outcome/ |
| 58 | (patient reported outcome measure* or patient outcome measure* or PROM* or POM* or outcome assessment*).mp. |
| 59 | or/53-58 |
| 60 | 4 and 52 |
| 61 | 59 and 60 |

Note: The final search strategies for the CTD and GAMT rapid literature reviews.

Embase (via OVID) and Medline (via OVID) were used to search for relevant papers.

Supplemental 4. Inclusion and exclusion criteria for the rapid literature reviews

| **Inclusion Criteria** |
|---|
| Subjects in the testing group of studies must be diagnosed with CTD or GAMT via genetic testing or biochemical confirmatory testing (e.g., brain creatine levels, creatine uptake studies, urine, plasma and CSF creatine or GAA levels, etc.). |
| Biomarkers, surrogate outcomes, medically important and patient/caregiver meaningful outcomes will be used as primary outcome measures. |
| Assessment tool/instrument (MRI, MRS, etc.) is described/outlined. |
| **Exclusion Criteria** |
| Full text not available online. |
| Experimental/animal studies. |
| Publication not available in English. |
| Published protocols. |
| Subjects diagnosed with other neurodevelopmental disorders in addition to CTD/GAMT (e.g., additional chromosomal abnormality). |

Note: Inclusion and exclusion criteria for CTD and GAMT rapid literature reviews.

Covidence was used for title/abstract and full text screening.

Supplemental 5. Focus group questions

1. What do you think are desirable outcomes that should be measured as part of a treatment study for children with CTD/GAMT? These could be important to you, your child, or both.
2. From the perspective of your child, what are the biggest challenges that they suffer from most?
3. From your perspective as a caregiver, what are the biggest challenges you face because of your child's creatine deficiency?
4. What do you want to see happen as an outcome of treatment?
5. What are your concerns or worries about a drug trial?
6. Are there outcomes that you feel are so ideal, you expect they are not achievable through a drug trial?

Supplemental 6. Mean Delphi survey outcome ratings.

| | Delphi 1 | | Delphi 2 | | Delphi 3 | |
|---|---|---|---|---|---|---|
| | Patient & Caregiver Mean Ratings (N) | Health Professional Mean Ratings (N) | Patient & Caregiver Mean Ratings (N) | Health Professional Mean Ratings (N) | Patient & Caregiver Mean Ratings (N) | Health Professional Mean Ratings (N) |
| Adaptive Functioning | 8 (90) | 8 (30) | 8 (83) | 8 (26) | 8 (63) | 8 (21) |
| Daily Living Skills | 8 (89) | 7 (29) | 8 (81) | 8 (27) | 8 (62) | 8 (21) |
| Cognitive Functioning | 9 (90) | 8 (30) | 9 (82) | 9 (27) | 9 (63) | 9 (21) |
| Emotional Dysregulation | 7 (90) | 6 (30) | 7 (83) | 6 (27) | 7 (63) | 6 (21) |
| MRS Brain Creatine | 9 (87) | 8 (27) | 9 (79) | 8 (27) | 8 (62) | 8 (21) |
| Seizures/ Convulsions | 8 (89) | 8 (29) | 8 (79) | 9 (27) | 8 (61) | 9 (21) |
| Serum/Plasma Guanidinoacetate (GAA) | 7 (77) | 8 (24) | 7 (77) | 8 (25) | 7 (58) | 8 (20) |
| Expressive Communication | 8 (90) | 8 (30) | 8 (82) | 8 (27) | 8 (63) | 8 (21) |
| Fine Motor Functions | 8 (87) | 6 (29) | 8 (79) | 7 (27) | 8 (60) | 7 (21) |
| MRI Brain, General | 8 (84) | 8 (28) | 8 (79) | 8 (27) | 8 (62) | 8 (21) |
| MRS Brain Guanidinoacetate | 8 (84) | 8 (23) | 8 (77) | 8 (25) | 8 (62) | 8 (19) |
| Serum/Plasma Creatine | 8 (85) | 7 (24) | 7 (80) | 8 (27) | 8 (60) | 8 (21) |
| Aggressive Behaviors | 7 (90) | 7 (30) | 7 (83) | 7 (27) | 7 (63) | 7 (21) |
| Caregiver Burden | 7 (88) | 7 (27) | 7 (79) | 8 (26) | 7 (60) | 8 (19) |
| EEG Epileptic Potentials | 8 (87) | 7 (28) | 8 (80) | 8 (27) | 8 (63) | 7 (21) |
| Executive Functioning | 8 (90) | 8 (30) | 8 (83) | 8 (26) | 8 (63) | 7 (20) |
| Independence | 8 (89) | 7 (29) | 8 (80) | 7 (27) | 8 (62) | 7 (21) |

| | | | | | | |
|---|---|---|---|---|---|---|
| Developmental Delay | 8 (89) | 8 (30) | 8 (82) | 9 (27) | 9 (63) | 9 (21) |
| Intellectual Disability | 8 (90) | 8 (30) | 9 (83) | 8 (27) | 9 (61) | 9 (21) |
| Life Expectancy | 7 (84) | 7 (26) | 7 (78) | 7 (27) | 7 (59) | 6 (20) |
| MRS Brain Phosphocreatine | 8 (86) | 8 (22) | 8 (79) | 8 (26) | 8 (61) | 9 (20) |
| Receptive Language | 8 (90) | 8 (30) | 8 (83) | 8 (27) | 8 (63) | 8 (21) |
| Treatment Access | 8 (87) | 7 (27) | 8 (80) | 8 (25) | 8 (61) | 8 (18) |
| Treatment Efficacy | 9 (88) | 8 (28) | 9 (80) | 9 (28) | 9 (61) | 9 (20) |
| Treatment Practicability | 8 (88) | 7 (29) | 8 (80) | 8 (27) | 8 (61) | 8 (20) |
| Treatment Safety | 9 (88) | 8 (29) | 9 (79) | 9 (27) | 9 (61) | 9 (20) |
| Treatment Tolerability | 8 (88) | 8 (29) | 8 (80) | 8 (27) | 8 (61) | 9 (20) |
| Coordination | 7 (87) | 6 (29) | 7 (79) | 7 (27) | 7 (60) | 7 (20) |
| EEG Abnormal Unspecific | 8 (88) | 6 (28) | 8 (80) | 6 (27) | 8 (62) | 7 (21) |
| Healthcare System Resource Availability | 7 (86) | 7 (27) | 7 (79) | 7 (26) | 7 (60) | 7 (19) |
| Low Muscle Tone, Hypotonia | 7 (86) | 7 (29) | 7 (79) | 7 (27) | 7 (60) | 7 (20) |
| MRI Brain Basal Ganglia | 8 (82) | 7 (27) | 7 (77) | 7 (27) | 7 (61) | 7 (20) |
| MRI Brain Atrophy | 8 (83) | 7 (29) | 8 (79) | 7 (27) | 7 (62) | 6 (21) |
| MRI Brain White Matter | 8 (83) | 7 (28) | 8 (79) | 7 (27) | 8 (61) | 7 (21) |
| MRI Cerebellar & Brainstem | 8 (83) | 7 (28) | 8 (77) | 7 (27) | 7 (61) | 6 (20) |
| Self-Protection | 7 (89) | 5 (29) | 7 (81) | 5 (27) | 7 (62) | 5 (21) |
| Sensory Processing Issues | 7 (89) | 6 (30) | 7 (82) | 6 (27) | 7 (63) | 6 (21) |
| Urine Creatine/Creatinine | 8 (88) | 7 (25) | 8 (81) | 8 (28) | 7 (62) | 8 (21) |

| | | | | | | |
|---|---|---|---|---|---|---|
| Walking Ability | 7 (86) | 7 (27) | 7 (79) | 7 (27) | 6 (60) | 7 (21) |
| Ataxia | 7 (84) | 7 (28) | 7 (77) | 7 (26) | 7 (59) | 7 (20) |
| Autism Spectrum Disorder (ASD) | 7 (89) | 7 (30) | 7 (81) | 8 (27) | 6 (63) | 7 (21) |
| QTc and Prolonged QTc | 7 (88) | 6 (27) | 7 (79) | 6 (27) | 7 (62) | 6 (21) |
| Attention Deficit/ Hyperactivity Disorder (ADHD) | 7 (89) | 7 (30) | 7 (82) | 6 (26) | 7 (63) | 6 (20) |
| CSF Creatine | 7 (80) | 6 (22) | 7 (77) | 6 (27) | 7 (58) | 6 (19) |
| CSF Guanidinoacetate (GAA) | 7 (74) | 6 (22) | 6 (71) | 6 (25) | 6 (55) | 5 (18) |
| Healthcare System Resource Use | 7 (87) | 7 (28) | 7 (78) | 7 (26) | 7 (60) | 7 (19) |
| High Muscle Tone, Spasticity | 6 (84) | 7 (28) | 6 (77) | 7 (27) | 6 (60) | 7 (20) |
| Sleep Disturbance | 6 (85) | 7 (28) | 6 (79) | 7 (27) | 6 (59) | 7 (20) |
| Social Relationships | 7 (89) | 6 (28) | 7 (81) | 6 (27) | 7 (62) | 6 (21) |
| Urine Guanidinoacetate (GAA) | 7 (80) | 7 (24) | 7 (74) | 7 (26) | 6 (60) | 6 (20) |
| Academic Achievement | 6 (90) | 6 (30) | 6 (82) | 5 (27) | - | - |
| Anxiety | 6 (90) | 6 (30) | 6 (82) | 6 (27) | - | - |
| Arrhythmia | 6 (89) | 6 (29) | 6 (81) | 6 (27) | - | - |
| Bladder Incontinence | 6 (87) | 5 (27) | 6 (81) | 5 (26) | - | - |
| Body Length | 5 (88) | 5 (29) | 5 (80) | 5 (27) | - | - |
| Breath-Holding | 6 (85) | 5 (26) | 6 (79) | 5 (26) | - | - |
| Cardiac Events | 7 (88) | 6 (29) | 7 (80) | 6 (27) | - | - |
| Cardiomyopathy | 6 (88) | 5 (28) | 6 (81) | 6 (27) | - | - |
| Constipation | 6 (88) | 6 (30) | 5 (83) | 5 (27) | - | - |
| Depression | 5 (88) | 6 (30) | 5 (82) | 6 (27) | - | - |

| | | | | | | |
|---|---|---|---|---|---|---|
| Dystonia | 6 (85) | 7 (29) | 6 (78) | 7 (27) | - | - |
| Educational Resource Use | 7 (86) | 7 (28) | 7 (79) | 7 (26) | - | - |
| Failure to Thrive | 7 (86) | 6 (28) | 6 (80) | 6 (27) | - | - |
| Feeding and Eating Problems | 6 (88) | 6 (30) | 6 (83) | 6 (27) | - | - |
| Frustration | 7 (90) | 5 (30) | 6 (83) | 5 (27) | - | - |
| Intolerance to Change | 6 (90) | 6 (30) | 6 (83) | 6 (27) | - | - |
| Low Energy | 6 (85) | 5 (29) | 6 (79) | 5 (27) | - | - |
| Muscle Weakness | 7 (86) | 6 (27) | 6 (78) | 6 (27) | - | - |
| Obsessive Compulsive Behavior | 6 (90) | 6 (30) | 6 (83) | 6 (27) | - | - |
| Oppositional Defiant Behavior | 7 (88) | 6 (30) | 6 (82) | 6 (27) | - | - |
| Pain | 6 (89) | 6 (30) | 6 (82) | 6 (27) | - | - |
| Physical Activity | 7 (87) | 5 (29) | 6 (80) | 5 (27) | - | - |
| Physical Endurance | 6 (86) | 5 (29) | 6 (79) | 5 (27) | - | - |
| Psychosis | 5 (85) | 6 (30) | 5 (81) | 6 (27) | - | - |
| Regression | 7 (90) | 7 (30) | 7 (82) | 7 (27) | - | - |
| Repetitive Behaviors | 7 (90) | 6 (30) | 6 (83) | 6 (27) | - | - |
| Restlessness/ Agitation | 7 (90) | 6 (30) | 7 (83) | 6 (27) | - | - |
| Self-Injury | 6 (88) | 7 (30) | 6 (82) | 7 (27) | - | - |
| Vision Impairment | 5 (88) | 6 (30) | 5 (82) | 5 (27) | - | - |
| Vomiting | 5 (87) | 5 (30) | 4 (82) | 5 (27) | - | - |
| Weight | 7 (88) | 6 (29) | 6 (80) | 6 (27) | - | - |
| Diarrhea | 5 (86) | 4 (30) | - | - | - | - |
| Gastroesophageal Reflux Disease (GERD) | 5 (87) | 5 (30) | - | - | - | - |

| | | | | | | |
|---|---|---|---|---|---|---|
| Hearing Impairment | 5 (90) | 6 (30) | - | - | - | - |
| Strabismus and Gaze Palsy | 5 (84) | 5 (30) | - | - | - | - |

Note: Mean ratings for each candidate outcome during the three Delphi survey rounds.

Supplemental 7. Consensus workshop voting results for the candidate outcomes.

| **Outcome** | **1-Definitely In n (%)** | **2-Maybe n (%)** | **3-Definitely Out n (%)** |
|---|---|---|---|
| Seizure/Convulsions | 24 (69) | 11 (31) | 0 (0) |
| Caregiver Burden | 2 (6) | 19 (54) | 14 (40) |
| Emotional Dysregulation | 21 (60) | 14 (40) | 0 (0) |
| Aggressive Behaviors | 1 (3) | 12 (34) | 22 (63) |
| Cognitive Functioning | 23 (66) | 12 (34) | 0 (0) |
| Intellectual & Developmental Disability | 5 (14) | 13 (37) | 17 (49) |
| Adaptive Functioning | 30 (86) | 5 (14) | 0 (0) |
| Expressive Language | 3 (9) | 2 (77) | 5 (14) |
| Receptive Language | 0 (0) | 22 (63) | 13 (37) |
| Independence | 1 (3) | 19 (54) | 15 (43) |
| Executive Functioning | 1 (3) | 14 (40) | 20 (57) |
| Fine Motor Functions | 0 (0) | 14 (40) | 21 (60) |
| MRS Brain Creatine | 29 (83) | 6 (17) | 0 (0) |
| MRS Brain Phosphocreatine | 2 (6) | 15 (43) | 18 (51) |
| MRI Brain General | 2 (6) | 15 (43) | 18 (51) |
| MRS Brain Guanidinoacetate | 0 (0) | 19 (54) | 16 (46) |
| EEG Epileptic Potentials | 0 (0) | 13 (37) | 22 (63) |
| Serum Plasma Creatine | 5 (14) | 10 (29) | 20 (57) |
| Serum Plasma Guanidinoacetate | 14 (40) | 8 (23) | 13 (37) |
| Life Expectancy | 0 (0) | 6 (17) | 29 (83) |

Note: There were 25 total voting consensus workshop participants. However, there were 35 voting responses as some health professionals voted twice, once for CTD and once for GAMT.
